# Development of a deep learning model for glomerulus detection and segmentation in kidney histopathology images

**DOI:** 10.64898/2026.10.19.26365514

**Authors:** Takeshi Tosaki, Takaya Sasaki, Yusuke Okabayashi, Kotaro Haruhara, Masahiro Okabe, Nobuo Tsuboi, Takashi Yokoo

## Abstract

**Background:** Estimation of nephron number, an indicator of renal reserve, requires quantitative assessment of glomerular number and size in kidney biopsy specimens. We aimed to develop an automated system for quantitative measurement of glomerular areas from clinical kidney biopsy images, across diverse kidney diseases.

**Methods:** We developed a two-stage deep learning system combining Faster R-CNN for glomerular detection in whole-slide images (WSIs) and DeepLabV3+ for segmentation. Of 114 kidney biopsies (228 WSIs), 104 were used for model development and 10 for independent final evaluation. Agreement was assessed using the F1-score for detection, the Dice coefficient for segmentation, and the intraclass correlation coefficient (ICC) for glomerular area measurement. Analysis was performed at 40× total magnification (2.331 μm/pixel) and validated against 400× images.

**Results:** In the independent evaluation set, the artificial intelligence (AI) model achieved a mean F1-score of 0.865 for detection, compared with 0.888 among experts. For segmentation, the Dice coefficient was 0.952 ± 0.015 and the per-pair ICC (2,1) for area measurement was 0.986 ± 0.013; both were comparable to inter-expert agreement. Bland–Altman analysis showed a mean bias of −344 μm² (95% limits of agreement, −1904 to 1216 μm²) between 40× and 400× measurements.

**Conclusion:** The two-stage model achieved glomerular segmentation and area measurement comparable to expert agreement in independent clinical data, with detection performance approaching inter-expert agreement, and provided reliable measurements at low magnification. This automated approach may facilitate large-scale quantitative kidney pathology and nephron number estimation from clinical kidney biopsies.

## Introduction

Nephron number is an important determinant of renal reserve and is associated with kidney function decline and progression of chronic kidney disease [1,2]. Although nephron number cannot be measured directly in living individuals, it can be estimated from kidney biopsy specimens using glomerular density and cortical volume [3]. Determining glomerular density requires morphometric analysis, including identification, counting, and area measurement of glomeruli in whole-slide images (WSIs).

However, manual identification and measurement of glomeruli are labor-intensive, time-consuming, and subject to interobserver variability, particularly when a large number of glomeruli must be analyzed [4–6]. Deep learning has recently been applied to automated glomerular detection and segmentation and has shown promising performance [7–11]. Nevertheless, pixel-level segmentation of entire WSIs at high resolution requires substantial computational resources, which limits its application in large-scale studies and routine pathology practice [12,13].

In this study, we developed a two-stage deep learning system for automated glomerular detection and segmentation using low-magnification WSIs. We aimed to evaluate its robustness using kidney biopsy specimens with diverse pathological backgrounds, assess its agreement with experts through independent evaluation by multiple nephrologists, and determine whether low-magnification analysis provides glomerular area measurements comparable to those obtained with conventional high-magnification analysis.

## Materials and Methods

### Study design and participants

This study was approved by the Institutional Review Board of The Jikei University School of Medicine (Approval No. 37-047[12684]) and conducted in accordance with the Declaration of Helsinki and the Declaration of Istanbul. The dataset consisted of 114 consecutive kidney biopsies performed at our institution between January 1, 2017, and December 31, 2017. The cohort comprised 102 native kidney biopsies and 12 kidney allograft biopsies, including 5 protocol biopsies obtained at the time of transplantation; all allografts were from living related donors. For each case, slides stained with periodic acid–Schiff (PAS) and hematoxylin and eosin (HE) were digitized as WSIs (total magnification, 40×), yielding a total of 228 WSIs. These two stains were selected because PAS is the standard stain for the histological assessment of kidney biopsy specimens, whereas HE is routinely available for kidney tissue obtained in other settings, including urological surgical specimens; a model that accepts both therefore has broader applicability. Images were acquired with a BX53 light microscope (Olympus, Tokyo, Japan), and adjacent fields covering the entire specimen were stitched into a single WSI using the multiple image alignment function of cellSens imaging software (Olympus).

The dataset was divided at the case level into a model development set (104 cases, 208 WSIs) and an independent evaluation set (10 cases, 20 WSIs). The development set was used for model training and validation, whereas the independent evaluation set was reserved exclusively for the final clinical evaluation. Details of image acquisition and dataset preparation are provided in **Supplementary Methods S1 and S2**.

### Artificial intelligence model

We developed a two-stage deep learning system for automated glomerular detection and segmentation using low-magnification WSIs (total magnification, 40×; **Fig. 1**). The first stage detected glomeruli within WSIs, and the second stage performed pixel-level segmentation of the detected glomeruli for morphometric analysis [14–17]. Glomerular profiles were annotated in six categories—non-sclerotic glomeruli (NSG), partial non-sclerotic glomeruli (pNSG), globally sclerotic glomeruli (GSG), partial globally sclerotic glomeruli (pGSG), Bowman’s capsule profiles without a visible tuft (CAPSULE), and partial Bowman’s capsule profiles (pCAPSULE)—and all six categories were used for detector training (**Supplementary Methods S3**). The distinction between non-sclerotic and globally sclerotic glomeruli is required for nephron number estimation [3]. Details of the model architecture, training procedures, data augmentation, and implementation are provided in **Supplementary Methods S4 and S5**.

**Fig. 1.**
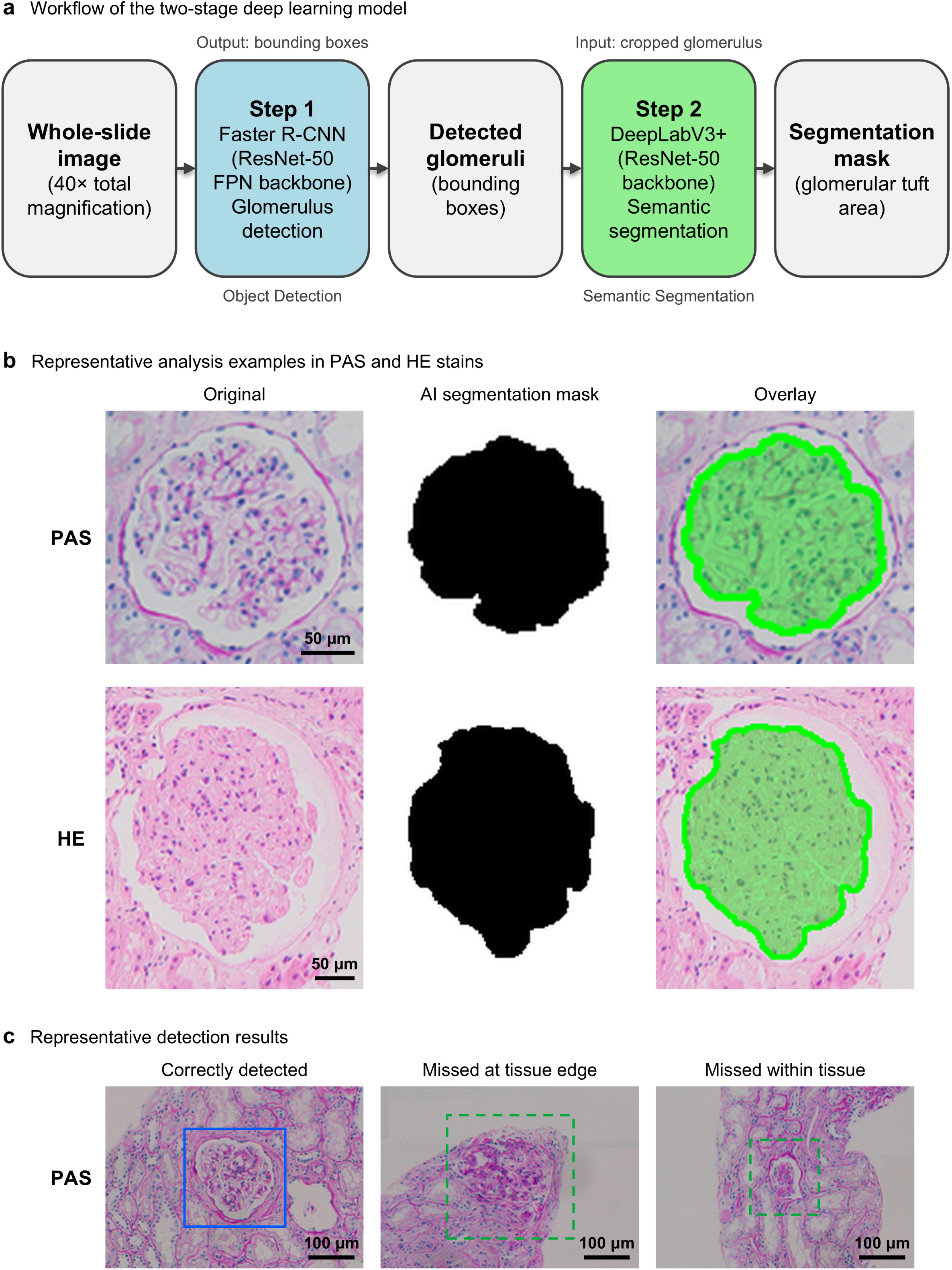
Construction of the AI analysis system and representative results. (a) Workflow of the two-stage deep learning model. WSIs at 40× total magnification are used as input. Step 1 (Faster R-CNN with a ResNet-50 feature pyramid network [FPN] backbone) detects glomeruli, and Step 2 (DeepLabV3+ with a ResNet-50 backbone) performs precise boundary extraction (segmentation). (b) Representative analysis examples of PAS- and HE-stained sections. From left: original histopathology image, AI-generated segmentation mask, and overlay of both. (c) Representative detection results from a PAS-stained whole-slide image in the independent evaluation set. Left, a glomerulus detected by the model (blue box); middle and right, glomeruli annotated by all four nephrologists but not detected by the model (green dashed boxes), located at the tissue edge (middle) and within the tissue (right). Their locations in the whole-slide image are shown in **Supplementary Figure 2a**. Scale bars, 50 μm (b) and 100 μm (c).

### Independent evaluation

To evaluate agreement with experts, four board-certified nephrologists who were not involved in model development independently evaluated the reserved evaluation set. Two validation tasks were performed. Task A assessed glomerular detection: each nephrologist marked all glomerular profiles within each WSI using the same six categories, and agreement was analyzed for non-sclerotic glomeruli with a visible tuft (NSG and pNSG; **Supplementary Methods S6**). Task B evaluated segmentation accuracy and glomerular area measurement using 100 non-sclerotic glomeruli with a visible glomerular tuft, randomly selected from the reference annotations of the evaluation set (**Supplementary Methods S6**).

### Validation of low-magnification analysis

To validate the use of low-magnification images for morphometric analysis, one nephrologist measured the glomerular area of a separate set of 100 glomeruli selected from the evaluation set using both low-magnification (40×) and conventional high-magnification (400×) images. The 40× images used in this comparison were generated by downsampling the 400× images by a factor of 10, so that the same glomeruli could be compared without image registration (**Supplementary Methods S7**).

### Statistical analysis

Because no single absolute ground truth exists for glomerular detection or segmentation, we adopted a pairwise agreement framework in which agreement between the artificial intelligence (AI) model and expert nephrologists was compared with the interobserver agreement among nephrologists.

For Task A, detection performance was evaluated using precision, recall, and F1-score based on an intersection-over-union (IoU) threshold of 0.5 [18]. For Task B, segmentation accuracy was assessed using the Dice similarity coefficient and IoU. Agreement for glomerular area measurement was evaluated using Pearson’s correlation coefficient, the intraclass correlation coefficient (ICC), and Bland–Altman analysis [19,20]. Agreement between measurements obtained at 40× and 400× magnification was evaluated using Bland–Altman analysis. Statistical analyses were performed using Python version 3.9 (**Supplementary Methods S8**).

## Results

### Study population and dataset

The dataset covered a wide spectrum of kidney pathologies, ranging from minor glomerular abnormalities to advanced global glomerulosclerosis (**Table 1**). A total of 228 PAS- and HE-stained WSIs from 114 kidney biopsies were included. The development set (104 cases) contained 3,801 glomeruli, whereas the independent evaluation set (10 cases) included 207 glomeruli according to the reference annotations. Compared with the development set, the independent evaluation set had a lower median estimated glomerular filtration rate (eGFR; 33.2 vs 52.9 mL/min/1.73 m²), indicating that the model was evaluated in cases with more advanced kidney disease, and a higher proportion of men (80.0% vs 55.8%). All five protocol biopsies obtained at the time of transplantation were assigned to the development set.

**Table 1.** Baseline characteristics of the study cohort (n = 114 biopsies).

| **Characteristic** | **Total<br>(n = 114)** |
|---|---|
| Age, years, mean ± SD | 50.3 ± 17.6 |
| Sex, male, n (%) | 66 (57.9%) |
| Serum creatinine, mg/dL, median [IQR] (n = 109) | 1.08 [0.77–1.78] |
| eGFR, mL/min/1.73 m<sup>2</sup>, median [IQR] (n = 109) | 50.3 [33.1–72.6] |
| Urinary protein-to-creatinine ratio, g/g, median [IQR] (n = 109) | 1.147 [0.351–3.470] |
| ***Biopsy type, n (%)*** |  |
| Native kidney | 102 (89.5%) |
| Kidney allograft | 12 (10.5%) |
| Protocol biopsy at transplantation | 5 (4.4%) |
| ***Underlying disease (native kidney biopsies), n (%)*** |  |
| IgA nephropathy | 32 (31.4%) |
| Focal global glomerular obsolescence | 14 (13.7%) |
| Focal segmental glomerulosclerosis (FSGS) | 9 (8.8%) |
| Membranous nephropathy (MN) | 7 (6.9%) |
| Lupus nephritis | 6 (5.9%) |
| Minimal change nephrotic syndrome | 6 (5.9%) |
| Glomerulonephritis, not otherwise specified | 4 (3.9%) |
| ANCA-associated vasculitis | 3 (2.9%) |
| Minor glomerular abnormalities | 3 (2.9%) |
| Other | 18 (17.6%) |

Continuous variables are shown as mean ± SD or median [interquartile range]. Underlying disease reflects the comprehensive clinical diagnosis where available; light-microscopy (HE/PAS) morphology was used when a comprehensive diagnosis was not otherwise documented. Serum creatinine, eGFR, and proteinuria were available in 109 of 114 biopsies; these values were not available for the five protocol biopsies obtained at the time of kidney transplantation (0–1 h). Protocol biopsies obtained at transplantation are a subset of the allograft biopsies. Percentages for underlying disease are calculated among the 102 native kidney biopsies; allograft biopsies are shown separately because they include protocol and follow-up biopsies performed irrespective of the original kidney disease. “Minor glomerular abnormalities” denotes biopsies without significant glomerular changes on light microscopy, as distinct from minimal change nephrotic syndrome, which was diagnosed in patients presenting with nephrotic syndrome.
Abbreviations: eGFR, estimated glomerular filtration rate; ANCA, antineutrophil cytoplasmic antibody.

### Task A: Glomerular detection performance

Pairwise agreement between the AI and each expert, together with inter-expert agreement, is shown in the agreement matrix (**Fig. 2a**). In the independent evaluation set (10 cases), the AI achieved F1-scores of ≥0.8 across all four AI–expert comparisons, with a mean F1-score of 0.865 (SD, 0.019). Although this was lower than the inter-expert agreement (mean F1-score, 0.888 [SD, 0.035]), the AI demonstrated a substantial level of agreement with individual experts. When the annotations of each expert were used as the reference, the precision of the AI ranged from 0.854 to 0.935 (mean, 0.902 [SD, 0.038]) and the recall from 0.797 to 0.867 (mean, 0.832 [SD, 0.030]).

**Fig. 2.**
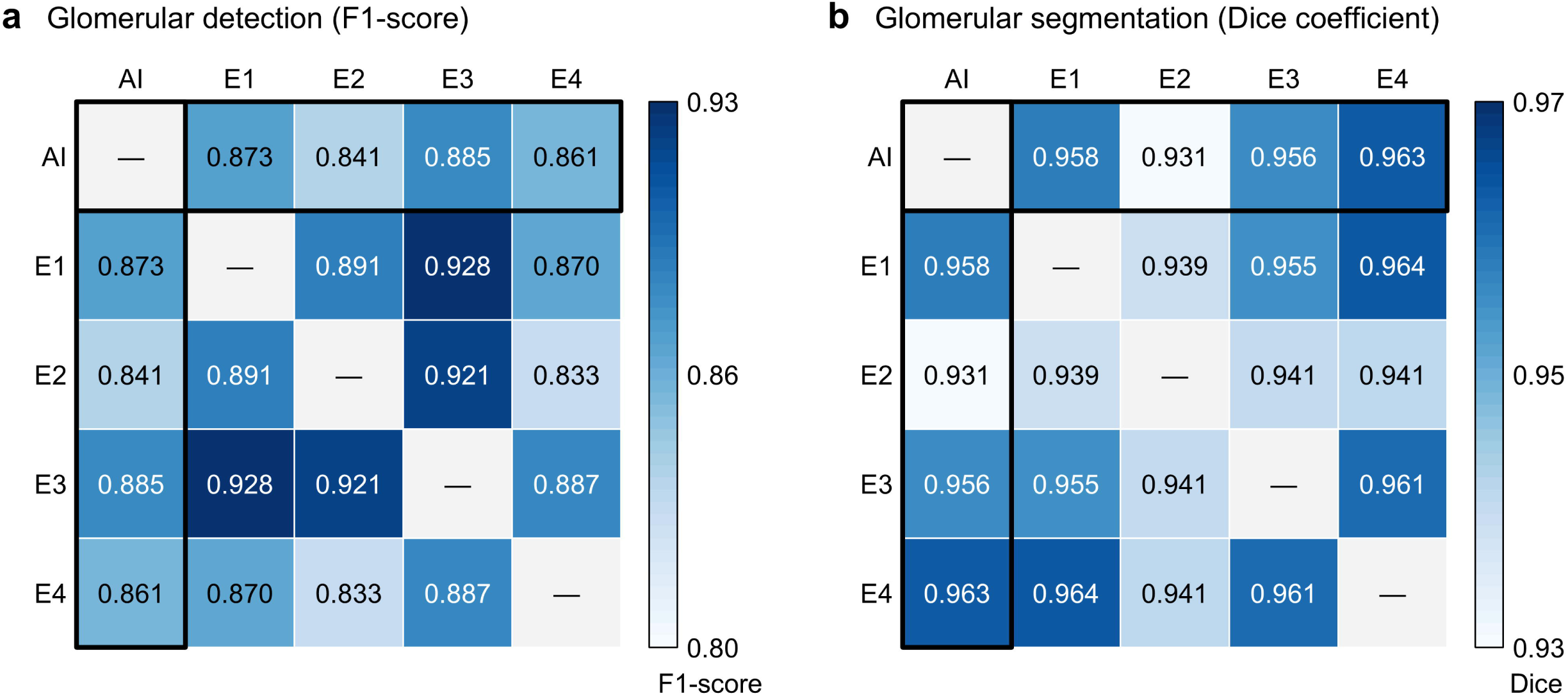
Pairwise agreement between the AI model and expert nephrologists. Heatmaps show pairwise agreement between the artificial intelligence (AI) model and four independent expert nephrologists (Evaluators 1–4). (a) Agreement for glomerular detection was assessed using the F1-score in the independent evaluation set. (b) Agreement for glomerular segmentation was assessed using the Dice similarity coefficient for 100 randomly selected glomeruli. Each cell represents the pairwise agreement between two evaluators, with darker shading indicating higher agreement. Black outlines highlight AI–expert comparisons, whereas the remaining off-diagonal cells represent inter-expert comparisons. Diagonal cells are omitted because they represent self-comparisons.

Pooled across the four comparisons, there were 78 false positives and 146 false negatives. The false positives arose from 38 model detections, 29 of which were annotated as non-sclerotic glomeruli by one to three of the four nephrologists. Of the nine detections regarded as false positives by all four nephrologists, eight had no corresponding annotation and one was annotated as a partial globally sclerotic glomerulus (**Supplementary Figure 2b**). Of the 146 false negatives, 117 (80.1%) had no corresponding detection (**Fig. 1c**), whereas 23 (15.8%) were detected but assigned to a category excluded from the detection analysis, most often GSG (n = 14).

### Task B: Glomerular segmentation and area measurement

Pairwise agreement between the AI and each expert, together with inter-expert agreement, is shown in the agreement matrix (**Fig. 2b**). In Task B (100 glomeruli), the AI achieved a Dice coefficient of 0.952 ± 0.015 and an IoU of 0.909 ± 0.026. For area measurement, the mean per-pair ICC (2,1) was 0.986 ± 0.013 (range, 0.968–0.995) for AI–expert comparisons, with a corresponding Pearson’s correlation coefficient of 0.992 ± 0.006. These values were comparable to those observed among expert–expert pairs (Dice, 0.950 ± 0.011; IoU, 0.906 ± 0.020; ICC (2,1), 0.978 ± 0.015), indicating a high level of agreement between the AI and individual experts that was comparable to inter-expert agreement. Bland–Altman analysis of glomerular area showed a mean bias of +456 μm² (AI minus expert; approximately 2% of the mean glomerular area) across AI–expert pairs (range, −439 to +1205 μm²), with limits of agreement comparable to those among expert–expert pairs.

### Validation of low-magnification morphometric analysis

Bland–Altman analysis of measurements obtained at 40× and 400× magnification by an expert showed a mean bias of −344 μm² (400× minus 40×; 95% limits of agreement, −1904 to 1216 μm²), corresponding to a relative bias of −2.4% (95% limits of agreement, −14.2 to 9.4%). The difference did not vary with glomerular size (regression of the difference on the mean area, P = 0.12), indicating a negligible systematic difference between magnifications. Representative images at 40× and 400× magnification are shown in **Fig. 3**. These findings demonstrate that the 40× magnification used in this study provides sufficient reliability for morphometric analysis.

**Fig. 3.**
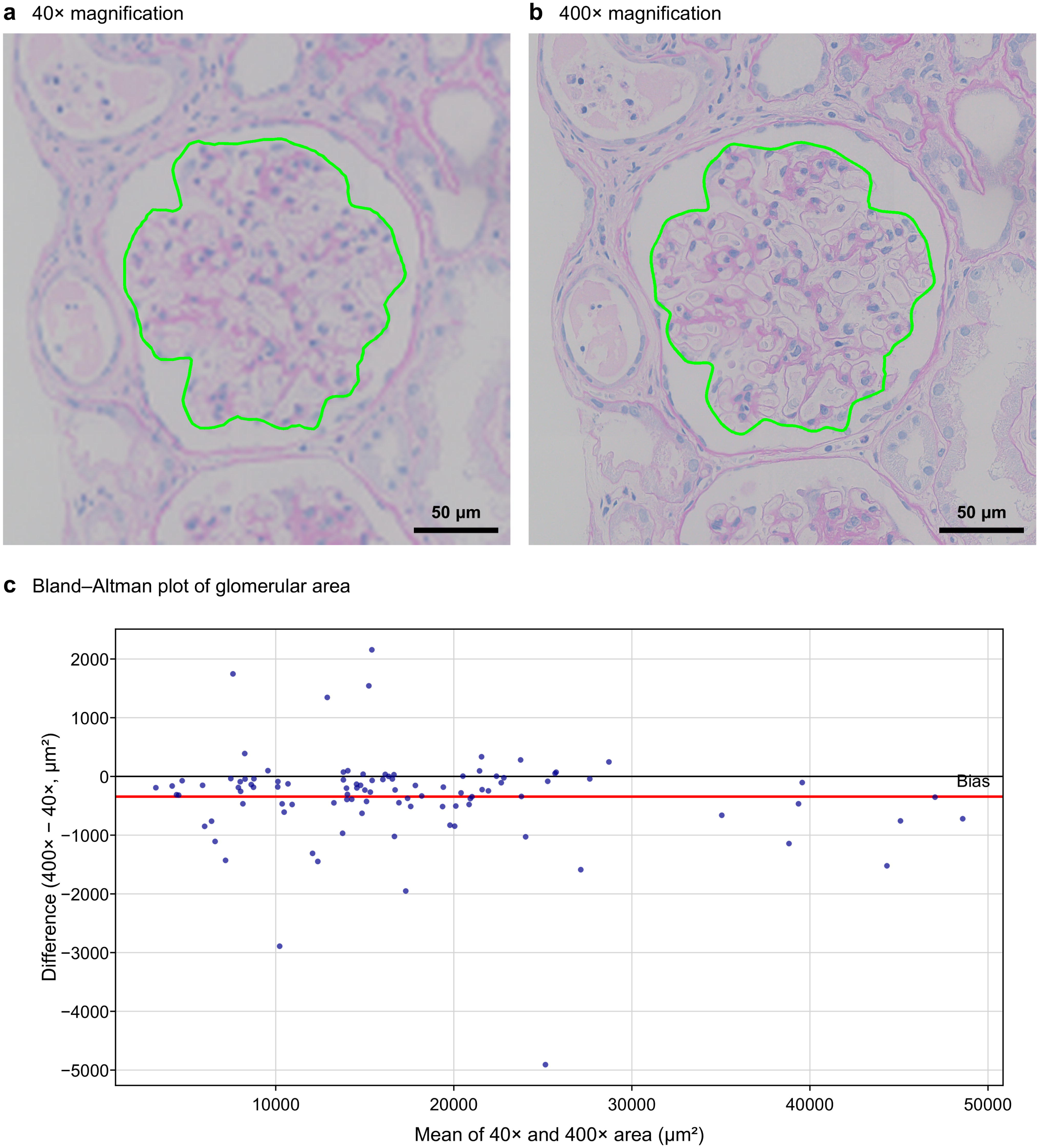
Validation of low-magnification morphometric analysis. (a) Representative image at 40× magnification. (b) Corresponding image at 400× magnification. Green outlines indicate the manually delineated glomerular tuft at each magnification. Scale bars, 50 μm. (c) Bland–Altman plot of area measurements obtained at 40× and 400× magnification (n = 100 glomeruli). The red line indicates the mean bias (400× minus 40×), and the black line indicates zero difference.

## Discussion

In this study, we developed and independently evaluated a two-stage deep learning system for automated glomerular detection and segmentation in kidney biopsy WSIs. The principal findings were that the model achieved glomerular segmentation and area measurements comparable to those of expert nephrologists, was evaluated across a heterogeneous spectrum of kidney pathologies, and enabled reliable morphometric assessment using low-magnification images. Importantly, the final evaluation was performed on cases completely independent of model development, using annotations from four independent experts, which allowed the performance of the model to be interpreted in the context of interobserver variability rather than against a single arbitrarily defined reference standard. These findings support the feasibility of using this approach for standardized quantitative morphometry in clinical kidney biopsy specimens.

Previous studies have demonstrated the feasibility of deep learning for glomerular detection, segmentation, and classification in kidney histopathology [4–6,8–11,13,21–23]. These studies have established the potential of artificial intelligence for automated assessment of renal tissue across different staining methods and pathological lesions. The present study extends these findings by focusing on a practical morphometric workflow for quantitative analysis of clinical kidney biopsy specimens. In particular, rather than evaluating model performance solely against a single reference annotation, we compared AI–expert agreement with inter-expert agreement using four independent evaluators. This approach is relevant because the identification and delineation of glomeruli, particularly globally sclerotic, incomplete, or poorly defined glomeruli, may themselves be subject to observer-dependent interpretation. The finding that AI–expert agreement for segmentation and area measurement was comparable to expert–expert agreement suggests that the model achieved a level of precision suitable for quantitative morphometric applications.

For glomerular detection, the mean F1-score of the AI model was approximately 0.87, slightly lower than the mean inter-expert agreement of approximately 0.89. Nevertheless, the model consistently achieved F1-scores above 0.8 across all AI–expert comparisons, indicating stable detection performance relative to each expert. Of the 38 detections counted as false positives against at least one expert, 29 were annotated as non-sclerotic glomeruli by other experts; discrepancies also arose in cases with advanced global glomerulosclerosis, where glomerular boundaries were poorly defined and interobserver agreement has been reported to be only moderate [5,12]. These findings suggest that at least part of the residual discrepancy reflects the inherent difficulty of defining such structures rather than model failure alone.

Another important finding was that glomerular area measurements obtained at 40× magnification showed negligible systematic differences from those obtained at 400× magnification. Although high-resolution images are advantageous for evaluating fine histological details, pixel-level analysis of entire WSIs at high resolution requires substantial computational resources [12,13]. Our findings suggest that low-magnification images provide sufficient resolution for glomerular morphometry while reducing the computational burden associated with large-scale image analysis. This feature may be particularly valuable for studies of nephron number, which require systematic identification, counting, and morphometric assessment of glomeruli across large numbers of biopsy specimens [3]. Automated extraction of glomerular number and area could substantially reduce the workload required for these analyses while improving reproducibility. More broadly, this approach may facilitate large-scale investigation of quantitative glomerular phenotypes, including glomerular density and size, and their relationships with nephron number, clinical characteristics, and kidney outcomes [24–27].

This study has several limitations. First, the model was developed and evaluated using data from a single center and a single imaging system. External validation using multicenter datasets, different scanners, and varying staining protocols is therefore needed to establish generalizability [25,28]. Second, the present study was limited to PAS- and HE-stained sections, and extension to other commonly used stains, including periodic acid–methenamine silver (PAM) and Masson trichrome, warrants further investigation [4,9]. Third, although glomerular identification and morphometry represent important components of nephron number estimation, comprehensive quantitative assessment of kidney tissue will ultimately require segmentation of additional structures, including tubules and the interstitium [10,21–23]. The accuracy of the six-category classification used to determine which glomeruli enter the morphometric analysis was not formally evaluated in the present study. Fourth, annotations in the development set were generated by a single evaluator and may therefore reflect annotator-specific tendencies; however, final model performance was evaluated against four independent experts who were not involved in model development. Fifth, the comparison between 40× and 400× magnification was performed by a single evaluator. Although the within-observer design allowed the effect of magnification to be assessed while minimizing interobserver variability, validation by multiple evaluators would strengthen this finding. Finally, partial glomeruli at tissue margins were included in the assessment of segmentation and area measurement. Their observed areas are systematically smaller than those of complete glomeruli; therefore, appropriate correction for partial glomeruli, as described in **Supplementary Methods S3** [3], is required when these measurements are applied to glomerular density or nephron number estimation.

In conclusion, the two-stage deep learning model developed in this study enabled automated glomerular detection, segmentation, and area measurement in kidney biopsy WSIs, with segmentation and glomerular area measurement comparable to inter-expert agreement in an independent evaluation set. Reliable area measurements were also obtained from low-magnification images, supporting a computationally efficient approach to quantitative glomerular morphometry. This system may provide a scalable and reproducible platform for large-scale kidney pathology research, including studies of glomerular morphology and nephron number estimation.

## Declarations

### Ethical approval

All procedures performed in the present study were in accordance with the ethical standards of the institutional research committee at which the study was conducted (Institutional Review Board of The Jikei University School of Medicine, approval number 37-047[12684]) and with the Declaration of Helsinki and the Declaration of Istanbul.

### Informed consent

Because of the retrospective nature of the study, the requirement for written informed consent was waived by the Institutional Review Board, and an opt-out approach was used. Information about the study was made publicly available, and patients were given the opportunity to decline participation.

### Clinical trial registration

Not applicable. This study was a retrospective observational study.

### Data availability

The data that support the findings of this study are available from the corresponding author upon reasonable request.

## Acknowledgements

The authors used ChatGPT (OpenAI) to improve the English language and readability of the manuscript. The authors reviewed and edited all AI-assisted content and take full responsibility for the final manuscript.

