## Supplementary Materials for "Development of a deep learning model for glomerulus detection and segmentation in kidney histopathology images"

### Supplementary Methods

#### S1. Whole-slide image preparation

Kidney biopsy specimens were routinely processed according to the standard pathological protocol at The Jikei University School of Medicine. For each case, slides stained with periodic acid–Schiff (PAS) and hematoxylin and eosin (HE) were prepared and imaged with a BX53 light microscope (Olympus, Tokyo, Japan). Adjacent fields covering the entire specimen were stitched into a single whole-slide image (WSI) using the multiple image alignment (MIA) function of cellSens imaging software (Olympus).

To improve computational efficiency while maintaining sufficient spatial information for glomerular analysis, all WSIs were acquired at a total magnification of 40× (2.331 μm/pixel). These low-magnification images were used for both model development and inference throughout the study.

#### S2. Dataset preparation

The dataset consisted of 114 kidney biopsies, comprising all native and allograft kidney biopsies performed at The Jikei University School of Medicine between January 1 and December 31, 2017. Each case included one PAS-stained and one HE-stained WSI, resulting in a total of 228 WSIs. Of the 114 cases, 102 were native kidney biopsies and 12 were kidney allograft biopsies, including 5 protocol biopsies obtained at the time of transplantation.

To avoid data leakage, all data splitting was performed at the case level so that images from the same patient were assigned exclusively to either the development or the evaluation set. The dataset was divided into two independent subsets: a model development set comprising 104 cases (208 WSIs) and an independent evaluation set comprising 10 cases (20 WSIs).

Within the development set (104 cases, 208 WSIs), images were randomly split into training and validation subsets in a 4:1 ratio using a fixed random seed. Case-level separation was applied between the development and independent evaluation sets, so that no image from an evaluation case was used at any stage of model development. The independent evaluation set was completely excluded from all stages of model development and was reserved exclusively for the final evaluation by independent nephrologists.

### S3. Annotation protocol

*Glomerular classification*

All glomeruli were manually annotated using rectangular bounding boxes for object detection and pixel-level masks for semantic segmentation. Six glomerular categories were defined according to their histological appearance:

- NSG (Non-sclerotic glomerulus): Glomerulus with a complete, non-sclerotic glomerular tuft.
- pNSG (Partial non-sclerotic glomerulus): Non-sclerotic glomerulus located at the tissue margin in which only part of the glomerular tuft is visible.
- GSG (Globally sclerotic glomerulus): Completely sclerotic glomerulus without a preserved glomerular tuft.
- pGSG (Partial globally sclerotic glomerulus): Partially visualized globally sclerotic glomerulus located at the tissue margin.
- CAPSULE: Bowman’s capsule profile without a visible glomerular tuft.
- pCAPSULE: Partial Bowman’s capsule profile located at the tissue margin.

Representative examples of each category are shown in **Supplementary Figure 1**.

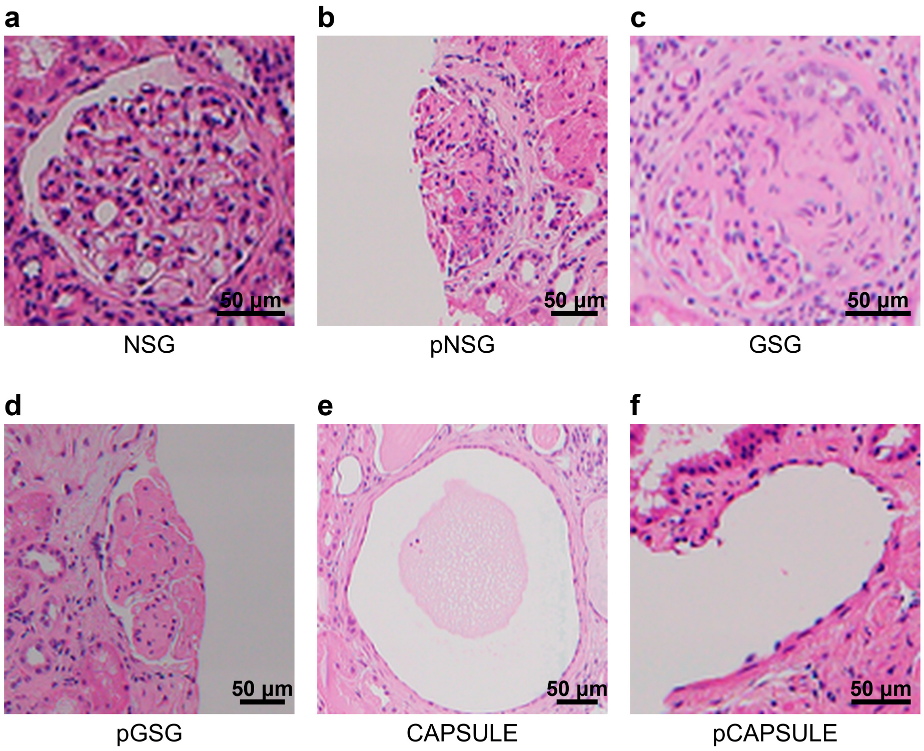


Supplementary Figure 1. Representative images of the six glomerular categories defined in the annotation protocol (**S3**). (a) NSG (non-sclerotic glomerulus). (b) pNSG (partial non-sclerotic glomerulus). (c) GSG (globally sclerotic glomerulus). (d) pGSG (partial globally sclerotic glomerulus). (e) CAPSULE (Bowman's capsule profile without a visible glomerular tuft). (f) pCAPSULE (partial Bowman's capsule profile). Hematoxylin and eosin (HE) stain.

*Reference annotations*

Reference annotations for both the model development set and the independent evaluation set were created by a nephrologist experienced in kidney morphometry (TT). To maintain annotation consistency, all annotations were repeatedly reviewed and self-cross-checked by the same annotator throughout the annotation process. These annotations served as the training reference during model development and as the sampling frame for Task B, and were not regarded as the definitive ground truth for final performance evaluation.

*Glomeruli included in morphometric analysis*

Morphometric analysis of glomerular area was performed only for glomeruli containing a visible glomerular tuft, namely NSG and pNSG. Globally sclerotic glomeruli (GSG and pGSG) were excluded because no preserved glomerular tuft was present, and CAPSULE and pCAPSULE were excluded because no glomerular tuft was visible.

For pNSG, the area of the visible glomerular tuft was measured directly without attempting geometric reconstruction of the missing portion outside the tissue section. This approach enabled direct comparison between AI-derived measurements and expert annotations even for glomeruli located at the tissue margin.

When glomerular density is subsequently used for nephron number estimation, partial glomeruli may be counted as 0.675 glomeruli according to the method proposed by Denic et al. [1]. However, this correction was not applied in the present validation study because the objective of Task B was to evaluate agreement in area measurement of the visible glomerular tuft rather than glomerular counting.

### S4. AI architecture

A two-stage deep learning framework was developed to perform automated glomerular detection and segmentation from low-magnification whole-slide images (WSIs). The workflow consisted of sequential object detection followed by semantic segmentation.

*Stage 1. Glomerular detection*

Glomeruli were first detected using Faster R-CNN with a ResNet-50 feature pyramid network (FPN) backbone implemented in PyTorch [2–5]. The detector identified candidate glomeruli and generated a rectangular bounding box for each detected object. All six glomerular categories (NSG, pNSG, GSG, pGSG, CAPSULE, and pCAPSULE) were included during detector training. For inference, each WSI was analyzed with a sliding window of 640 × 640 pixels with 10% overlap. Detections with a confidence score of ≥0.5 were retained, and overlapping detections were merged by non-maximum suppression (IoU threshold, 0.1) applied across all categories, so that each glomerulus was assigned a single category.

*Stage 2. Glomerular segmentation*

Each detected glomerulus was subsequently cropped from the WSI and processed using a DeepLabV3+ semantic segmentation network with a ResNet-50 backbone [2,6]. The segmentation model generated pixel-level masks of the glomerular tuft, which were subsequently used for morphometric analysis, including glomerular area measurement.

The two-stage framework reduced computational cost by restricting pixel-level segmentation to glomerular regions detected during the first stage rather than processing the entire WSI.

**S5. Model training**

*Stage 1. Detection model*

The Faster R-CNN detection model was trained using the AdamW optimizer with an initial learning rate of 0.0001 and a batch size of four. The maximum number of training epochs was set to 150. To improve generalizability, online data augmentation was applied during training using random horizontal and vertical flipping together with color transformations. Training was terminated using an early stopping strategy when the validation loss failed to improve for 15 consecutive epochs.

*Stage 2. Segmentation model*

The DeepLabV3+ segmentation model was trained using Dice loss and the Adam optimizer with an initial learning rate of 0.001. Data augmentation was performed using the Albumentations library [7] and included random horizontal and vertical flipping, rotation, ShiftScaleRotate, and blur transformations. Early stopping was applied using the validation loss with a patience of 20 epochs.

*Computational environment*

Model development and training were performed on a workstation equipped with an NVIDIA GeForce RTX 4070 Ti SUPER graphics processing unit (GPU). Additional implementation details, including software versions, are available from the corresponding author upon reasonable request. Both models were implemented using the PyTorch deep learning framework.

### S6. Independent evaluation

*Independent expert evaluation*

To evaluate agreement of the proposed model with experts, an independent evaluation was conducted using the reserved evaluation set (10 cases, 20 WSIs). Four board-certified nephrologists who had not participated in model development or in the creation of the reference annotations independently evaluated the WSIs.

Because no single observer can be regarded as the absolute ground truth for glomerular detection or segmentation, the independent evaluation was designed to compare the performance of the AI model with the level of agreement observed among expert nephrologists.

*Task A. Glomerular detection*

For each WSI, all glomerular profiles, including globally sclerotic and partial glomeruli and Bowman's capsule profiles (all six categories defined in **S3**), were independently identified by each nephrologist using bounding boxes. Detection agreement was then calculated for non-sclerotic glomeruli with a visible tuft (NSG and pNSG); globally sclerotic glomeruli and Bowman's capsule profiles were annotated but excluded from the analysis, because their identification is not required for glomerular density estimation. Detection performance was evaluated by comparing every pair of observers, including AI–expert and expert–expert pairs. Representative detection results are shown in **Supplementary Figure 2**.

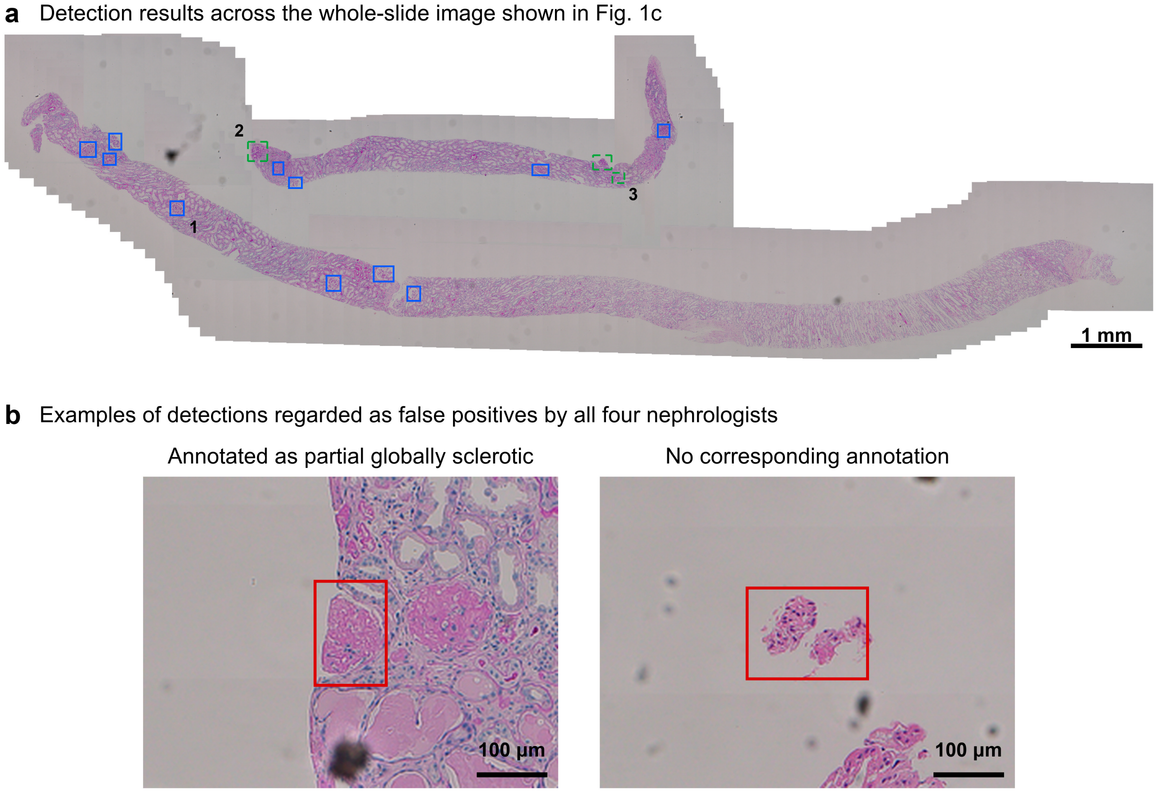


Supplementary Figure 2. Representative detection results in the independent evaluation set. (a) Detection results across the periodic acid–Schiff (PAS)-stained whole-slide image shown in **Fig. 1c**. Blue boxes indicate glomeruli detected by the model, and green dashed boxes indicate glomeruli annotated by all four nephrologists but not detected by the model. Glomeruli annotated by only some of the nephrologists are not shown, and no false-positive detection occurred in this whole-slide image. Numbers 1–3 indicate the glomeruli shown in

**Fig. 1c** (left, middle, and right, respectively). (b) Examples of detections regarded as false positives by all four nephrologists (red boxes): a partial glomerulus detected as non-sclerotic by the model but annotated as partial globally sclerotic by all four nephrologists (left; PAS stain) and a small tissue fragment detached from the section without any corresponding annotation (right; HE stain). Scale bars, 1 mm (a) and 100 μm (b).

*Task B. Glomerular segmentation and area measurement*

For segmentation analysis, 100 non-sclerotic glomeruli (NSG or pNSG) were randomly selected from the reference annotations of the independent evaluation set as a fixed evaluation set. Each nephrologist manually delineated the glomerular tuft at the pixel level. Glomerular area was calculated as the total number of pixels contained within each segmentation mask. These measurements were subsequently used for agreement analyses between the AI model and the nephrologists.

**S7. Validation of low-magnification analysis**

To evaluate whether low-magnification images provide morphometric measurements comparable to those obtained from conventional high-magnification images, one experienced nephrologist measured the glomerular area of a separate set of 100 glomeruli selected from the evaluation set using both low-magnification (40×) and high-magnification (400×) images. The 40× images used in this comparison were generated by downsampling the 400× images by a factor of 10, so that the same glomeruli could be compared without image registration.

**S8. Statistical analysis**

Since no absolute reference standard exists for glomerular detection or segmentation, we adopted a pairwise agreement framework rather than selecting a single observer as the ground truth.

Agreement was calculated for every unique observer pair among the AI model and the four nephrologists, yielding ten pairwise comparisons (four AI–expert pairs and six expert–expert pairs). The performance of the AI model was considered comparable to expert performance when the mean agreement between the AI and nephrologists fell within the range of agreement observed among nephrologists.

Detection accuracy was evaluated using an intersection-over-union (IoU) threshold of 0.5. Precision, recall, and F1-score were calculated for each observer pair by pooling true positives, false positives, and false negatives across all WSIs.

Segmentation performance was evaluated using the Dice similarity coefficient (DSC) and IoU for each observer pair. Results are presented as the mean ± standard deviation.

Agreement of glomerular area measurements was evaluated using three complementary statistical methods. First, Pearson's correlation coefficient (r) and the coefficient of determination ($R^2$) were used to assess the linear relationship between paired measurements. Second, the intraclass correlation coefficient ICC (2,1), based on a two-way random-effects model with absolute agreement, was calculated to quantify measurement agreement. Third, Bland–Altman analysis was performed to estimate the mean bias and the 95% limits of agreement (mean bias ± 1.96 standard deviations) and to evaluate systematic and proportional measurement errors. Scatter plots with regression lines and Bland–Altman plots were generated for each observer pair.

Agreement between measurements obtained at 40× and 400× magnification was evaluated using Bland–Altman analysis, expressed both as absolute differences and as percentages of the mean of the paired measurements. Proportional bias was assessed by regressing the difference on the mean of the paired measurements.

All statistical analyses were performed using Python version 3.9 (NumPy, SciPy, Pandas, Statsmodels, Matplotlib, and OpenCV).
